# ROX Index Predicts Intubation in Patients with COVID-19 Pneumonia and Moderate to Severe Hypoxemic Respiratory Failure Receiving High Flow Nasal Therapy

**DOI:** 10.1101/2020.06.30.20143867

**Authors:** Maulin Patel, Junad Chowdhury, Nicole Mills, Robert Marron, Andrew Gangemi, Zachariah Dorey-Stein, Ibraheem Yousef, Matthew Zheng, Lauren Tragesser, Julie Giurintano, Rohit Gupta, Parth Rali, Gilbert D’Alonzo, Huaqing Zhao, Nicole Patlakh, Nathaniel Marchetti, Gerard J. Criner, Matthew Gordon, for the Temple University COVID-19 Research Group

**Author notes:** see supplement (for the Temple University COVID-19 Research Group). **Contact Information:** Maulin Patel, MD, Temple University Hospital, 7^th^ floor Parkinson Pavilion, 3401 N Broad St, Philadelphia, PA 19140. Contributions: Maulin Patel will be the corresponding author and guarantor for the manuscript. Maulin Patel, Matthew Gordon, Junad Chowdhury and Gerard J Criner formulated the overall study design. Huaqing Zhao, Nicole Patlakh, Maulin Patel, Andrew Gangemi, Robert Marron, Junad Chowdhury, Nicole Mills, Zachariah Dorey-Stein, Ibraheem Yousef, Lauren Tragesser, Julie Giurintano assisted in data collection, consolidation and analysis. Maulin Patel, Junad Chowdhury, Parth Rali, Rohit Gupta, Gilbert D’Alonzo and Matthew Gordon drafted the manuscript. Gerard J Criner and Matthew Gordon revised and reviewed the Manuscript.

## Abstract

Use of high flow nasal therapy (HFNT) to treat COVID-19 pneumonia has been greatly debated around the world due to concern for increased healthcare worker transmission and delays in invasive mechanical Ventilation (IMV).

**Methods:** A retrospective analysis of consecutive patients admitted to Temple University Hospital in Philadelphia, Pennsylvania, from March 10, 2020, to May 17, 2020 with moderate to severe respiratory failure treated with High Flow nasal therapy (HFNT). HFNT patients were divided into two groups: HFNT only and HFNT progressed to IMV. The primary outcome was the ability of the ROX index to predict the need of IMV.

**Results:** Of the 837 patients with COVID-19, 129 met inclusion criteria. The mean age was 60.8 (±13.6) years, BMI 32.6 (±8), 58 (45 %) were female, 72 (55.8%) were African American, 40 (31%) Hispanic. 48 (37.2%) were smokers. Mean time to intubation was 2.5 days (± 3.3). ROX index of less than 5 at HFNT initiation was predictive of progression to IMV (OR = 2.137, p = 0,052). Any decrease in ROX index after HFNT initiation was predictive of intubation (OR= 14.67, p <0.0001). **Δ**ROX (<=0 versus >0), peak D-dimer >4000 and admission GFR < 60 ml/min were very strongly predictive of need for IMV (ROC = 0.86, p=). Mortality was 11.2% in HFNT only group versus 47.5% in the HFNT progressed to IMV group (p,0.0001). Mortality and need for pulmonary vasodilators were higher in the HNFT progressed to IMV group.

**Conclusion:** ROX index is a valuable, noninvasive tool to evaluate patients with moderate to severe hypoxemic respiratory failure in COVID-19 treated with HFNT. ROX helps predicts need for IMV and thus limiting morbidity and mortality associated with IMV.

## Introduction

December of 2019 was marked by a cluster of acute respiratory illnesses now known as Coronavirus disease 2019 (COVID-19) resulting from severe acute respiratory syndrome-coronavirus-2 (SARS-CoV-2). The virus has infected more than 8.7 million people worldwide with more than 460,000 reported deaths resulting in a worldwide healthcare crisis.^1,2^ The majority of morbidity from COVID-19 arises from severe hypoxemic respiratory failure. As the worldwide pandemic spreads to the farthest reaches of the globe, healthcare centers have been overwhelmed, quickly exhausting their supply of ventilators and personnel trained to manage them. There has been significant controversy regarding the optimal mode of respiratory support to treat COVID-19 associated hypoxemic respiratory failure.

The timing and adequacy of non-invasive forms of oxygen support (i.e. high flow nasal therapy, simple face mask, etc.) versus invasive mechanical ventilation (IMV) is not known. IMV has been associated with significant morbidity and mortality. In some case series, a mortality rate greater than 90% has been reported.^3-6^ Case series from China, Italy and New York had intubation rates ranging from 20.2% to 88%.^4,6-9^ Early utilization of IMV has been greatly influenced by concerns of viral aerosolization and subsequently health care transmission through the use of non-invasive forms of oxygen support.^10^ In addition, rapid progression of hypoxemic respiratory failure from mild dyspnea to ARDS within 48-72 hours were noted in early studies.^9,11^ Consequently, some centers preemptively intubate patients with oxygen requirements as low as 6 liters-per-minute (LPM) of oxygen via nasal cannula for prolonged periods.^3^

High flow nasal therapy (HFNT), in contrast to IMV, is a non-invasive oxygen system that delivers humidified air-oxygen blends and a titratable fraction of inspired oxygen (F_I_O_2_) as high as 60 LPM and 100% F_I_O_2_ respectively. Despite proven efficacy in other disease processes, the utilization of HFNT has been limited and its use has not been widely recommended for use in patients with COVID-19 pneumonia and hypoxemic respiratory failure. Limitations to adoption include concerns about rapid progression of disease as well as fearing aerosolization of the COVID-19 virus resulting in increased transmission to healthcare providers.^12-14^

However, HFNT has been successfully used in severe viral respiratory illnesses including influenza A and H1N1.^15^ HFNT reduces the need for invasive mechanical ventilation rates compared to other modalities, with some studies also showing reduced 90-day mortality.^16-19^ By decreasing the incidence of invasive ventilation, HFNT has the potential to decrease complications associated with IMV such as the incidence of ventilator-associated pneumonia (VAP). When compared with noninvasive ventilation (NIV) and conventional oxygen therapy, the use of HFNT has also shown to reduce rates of reintubation due to post-extubation respiratory failure and has much better tolerability than NIV.^20,21^ The Surviving Sepsis Guidelines for COVID-19 also recommends using HFNT in patients with acute hypoxemic respiratory failure due to COVID-19.^22^

The ROX index, defined as the ratio of oxygen saturation as measured by pulse oximetry (SpO2)/FiO2 to respiratory rate (RR) in breaths per minute, is a validated measurement that predicts outcomes when using HFNT to treat hypoxemic respiratory failure. A ROX index < 4.88 after 12 hours predicts the need for IMV in patients with pneumonia.^23^

Herein we analyze the utility of the ROX index to predict the need and timing for IMV in a retrospective analysis of 129 patients with COVID-19 associated with moderate to severe hypoxemic respiratory failure treated with HFNT. In addition, mortality, rates of intubation, length of stay (LOS) and rates of nosocomial infections in our cohort treated with HFNT were also reported.

## Methods

The study was approved by the Temple University Institutional Review Board (TU-IRB protocol number: 27051). A waiver of consent was granted due to the acknowledged minimal risk to the patients.

A retrospective analysis of 1397 consecutive patients admitted to Temple University Hospital in Philadelphia, Pennsylvania, from March 10, 2020, to May 17, 202 was performed. Initial screening included all patients who had tested positive for COVID-19 using nasopharyngeal real time reverse transcriptase PCR (RT-PCR) or had high clinical suspicion based on high-resolution computerized tomography (HRCT) of the chest (typical peripheral nodular or ground glass opacities without alternative cause)^24^ with typical inflammatory biomarker profile and a suggestive clinical history.

All patients with moderate to severe hypoxemic respiratory failure who were treated with HFNT at any point during the hospitalization were included in the study. Moderate and severe hypoxemic respiratory failure was defined as hypoxemia requiring more than 6 L/min of oxygen via nasal cannula. Absence of HFNT during hospitalization was an exclusion criterion.

### Laboratory data

Demographics including age, sex, comorbidities, body mass index (BMI), and smoking status (current smoker, non-smoker) were collected. In addition, laboratory biomarkers on admission including complete blood count (CBC) with differential, ferritin, fibrinogen, lactate dehydrogenase (LDH), D-dimer, and C-reactive protein (CRP) were analyzed.

### Respiratory Metrics

Respiratory metrics at the initiation of HFNT included respiratory rate (RR), pulse oximetry, and fraction of inspired oxygen (F_I_O_2_). The same parameters were collected at days 1, 2, 3 and 5 post-initiation of HFNT. Parameters were recorded at the lowest F_I_O_2_ and highest pulse oximetry reported for the day. For patients who required IMV prior to the conclusion of data collection, respiratory parameters on the day of intubation were reported. Days on HFNT therapy, time to intubation (in days), average flow rate on HFNT, and the presence of hospital acquired pneumonia (HAP)/Ventilator associated Pneumonia (VAP) were also reported.

### Respiratory therapy

HFNT was provided with a humidified air-oxygen blender starting at 35 LPM with the fraction of inspired oxygen (F_I_O_2_) adjusted to maintain oxygen saturations ≥ 92%; further adjustments were made based on patients’ tolerance and goals of oxygenation. The initial temperature for the high flow setup was 37° C and was titrated between 34-37° C for patient comfort. As an institutional policy, HFNT was preferred over IMV and was maintained indefinitely as long as oxygenation, ventilation, and work of breathing parameters were acceptable. Data on initial oxygenation support included flow of air-oxygen blend in LPM and F_I_O_2_. The decision to switch to NIV or IMV was at the discretion of the clinical care team. Once on IMV, patients were assessed daily for appropriateness of spontaneous breathing trials (SBT) and spontaneous awakening trials (SAT) for extubation per standard guidelines. Patients were extubated following a SBT of 15 - 30 minutes on either pressure support ventilation (PSV) of 5cmH_2_O or T-piece with a viral filter based on clinical judgement and rapid shallow breathing index.

### Institutional Approach to COVID-19 Directed Therapies

All patients admitted due to respiratory symptoms and classic radiographic evidence of COVID-19 pneumonia were admitted to a specialized hospital unit and given antibiotic therapy for community acquired pneumonia and systemic steroids with methylprednisolone at 0.5 to 1.0 m /kg for at least 5-10 days. Patients with RT-PCR positive swabs were also screened for eligibility for randomized controlled trials at our institution which included sarilumab (Regeneron Pharmaceuticals; NCT04315298), remdesivir (Gilead Sciences; NCT04292730 and NCT04292899), gimsilumab (Kinevant

Sciences: NCT04351243), and convalescent plasma (Mayo Clinic; NCT04338360). Those with significant disease, but ineligible for clinical trials were treated with compassionate use of anakinra, tocilizumab or etoposide based on institutional care pathways. Other therapies included high-dose corticosteroids (defined as a minimum daily 125 mg of methylprednisolone and above, regardless of bolus frequency), intravenous immunoglobulin (IVIG), and hydroxychloroquine (HCQ). Therapies were offered based on clinical severity, radiographic burden, and/or presence of cytokine storm as evidenced by inflammatory markers. The decision to select from these choices was made by the same institutional multidisciplinary team including pulmonologists and rheumatologists.

### Outcomes

The primary outcome was the ability of the ROX index to predict the need of IMV. Secondary outcomes include mortality, hospital length of stay (LOS) and hospital/ventilator acquired pneumonia. Hospital and ventilator acquired pneumonia was defined based on the presence of sputum positivity and treatment with antibiotics.

Our patients were divided into two groups: 1) HFNT support as a bridge to recovery (HFNT group) and 2) HFNT with progression to IMV (i.e., intubation group) for analysis. Comparison was made between demographics, baseline laboratory values, and outcomes within the two groups. Changes in ROX index and concomitant changes in clinical parameters of heart rate (HR) were also analyzed.

A multivariable prediction model for intubation for our cohort based on the above parameters was created. ROX index, comorbidities, clinical and laboratory data were used to identify parameters that could predict the need for intubation. A ROC curve was going to be generated to determine accuracy of the model.

### Statistical methods

Continuous variables are presented as means (± standard deviation), and categorical variables as numbers and Frequency (percentages). Continuous variables were compared with the use of the two-sample t-test or paired t-test for categorical variables with the use of the Pearson chi-square test. Laboratory data were nonparametric and compared using Wilcox Rank-Sum test. Kaplan-Meier analysis was estimated for survival and compared by log-rank test.

To build a predictive model of the intubation, multivariable logistic regression was performed to determine the adjusted associations of the variables with intubation. The initial model included all the variables associated with intubation in univariate analyses for p<0.1. The final model that optimized the balance of the fewest variables with good predictive performance. Assessment of model performance was based on discrimination and calibration. Discrimination was evaluated using the C-statistic, which represents the area under the receiver operating characteristic (ROC) curve, where higher values represent better discrimination. Calibration was assessed by the Hosmer-Lemeshow test, where a p-value greater than 0.05 indicates adequate calibration.

All statistical tests were two-tailed, and P values of less than 0.05 were considered to indicate statistical significance. All statistical analyses were performed with the use of Stata 14.0 (StataCorp LP, College Station, TX).

## Results

### Patient population

1,397 patients admitted to Temple University Hospital between March 10, 2020, and May 17, 2020 were screened. Of these, 837 patients had tested positive for COVID-19 by nasopharyngeal RT-PCR or were treated for high clinical suspicion based on typical CT imaging and inflammatory biomarker profile. 388 patients had hypoxemic respiratory failure. 129 (15.4%) patients met our inclusion criteria of being on High flow nasal therapy (HFNT) with moderate to severe hypoxemic respiratory failure (Figure 1).

**Figure 1:**
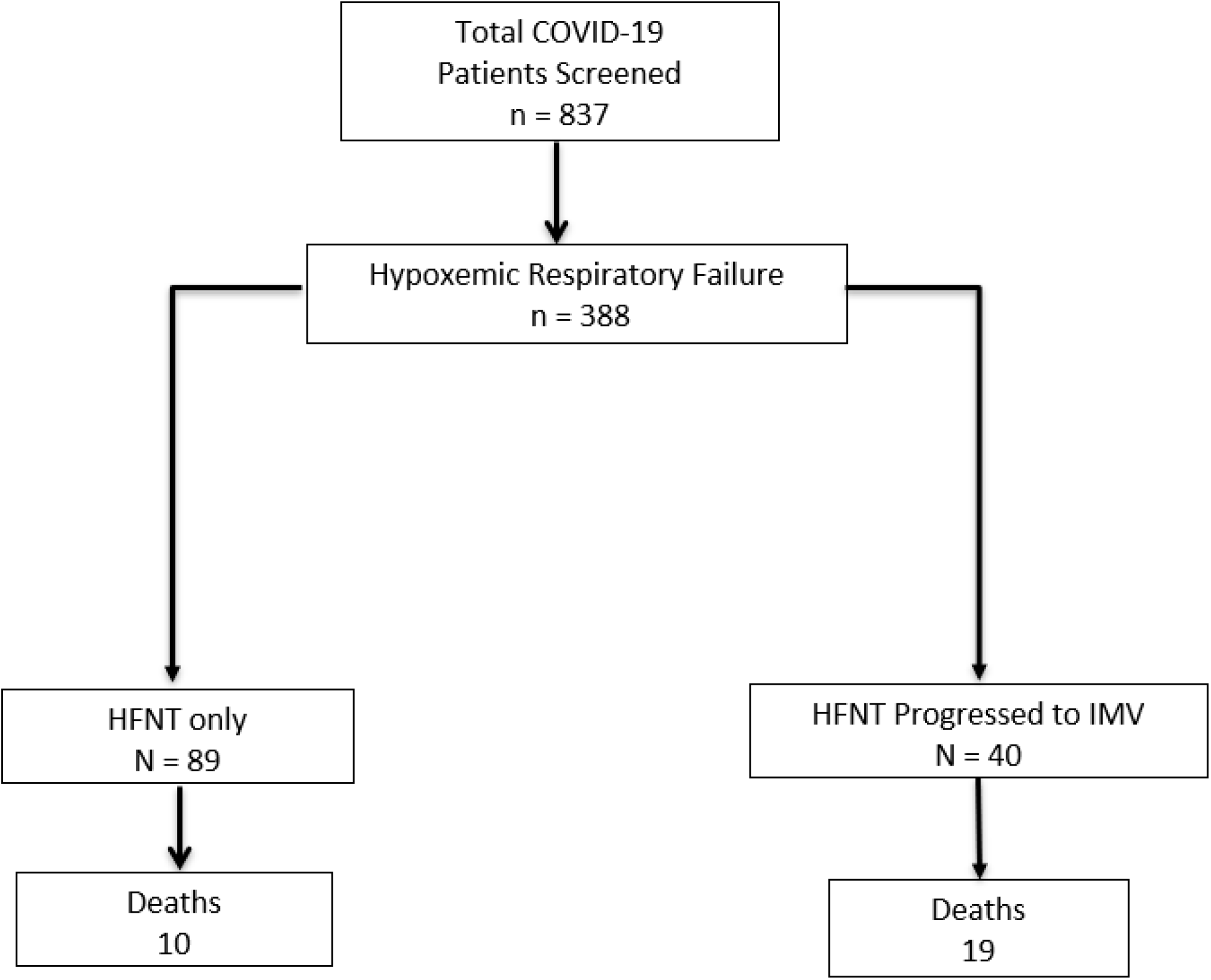
Consort Diagram for our screening.

### Demographics

The mean age was 60.8 (±13.6) years, BMI 32.6 (8), 58 (45 %) were female, 72 (55.8%) were African American, 40 (31%) Hispanic. 48 (37.2%) were smokers. The major comorbidities reported (in descending incidence) were hypertension, diabetes, lung disease, heart disease, chronic kidney disease (CKD), malignancy and psychiatric illness (Table 1). There were no differences in age, BMI and gender between the groups. There were more smokers in the intubation group at 55% compared to 29.2%. There was a trend towards a higher incidence of lung disease, CKD, malignancy and psychiatric disorders in the intubation group.

**Table 1:**
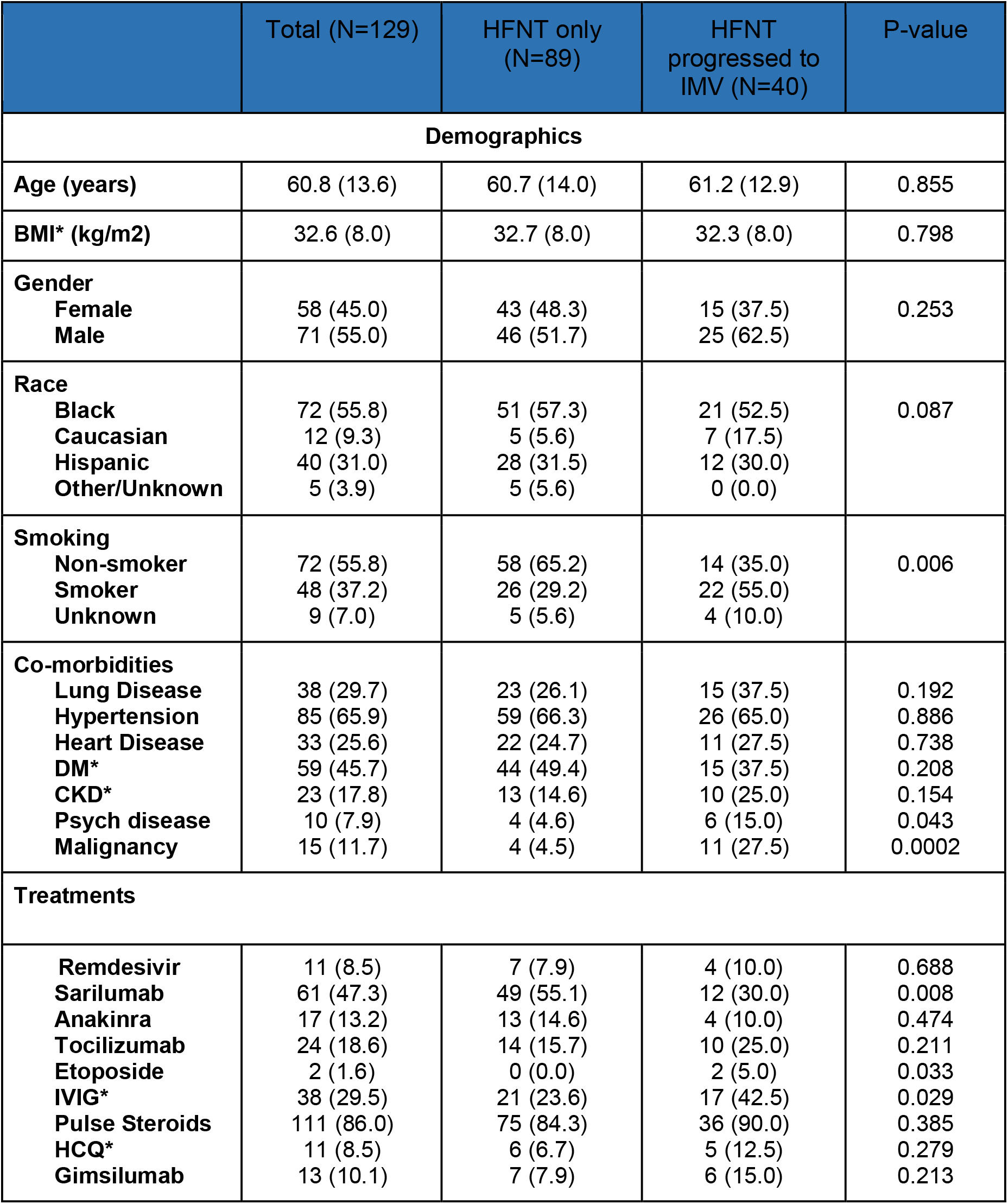
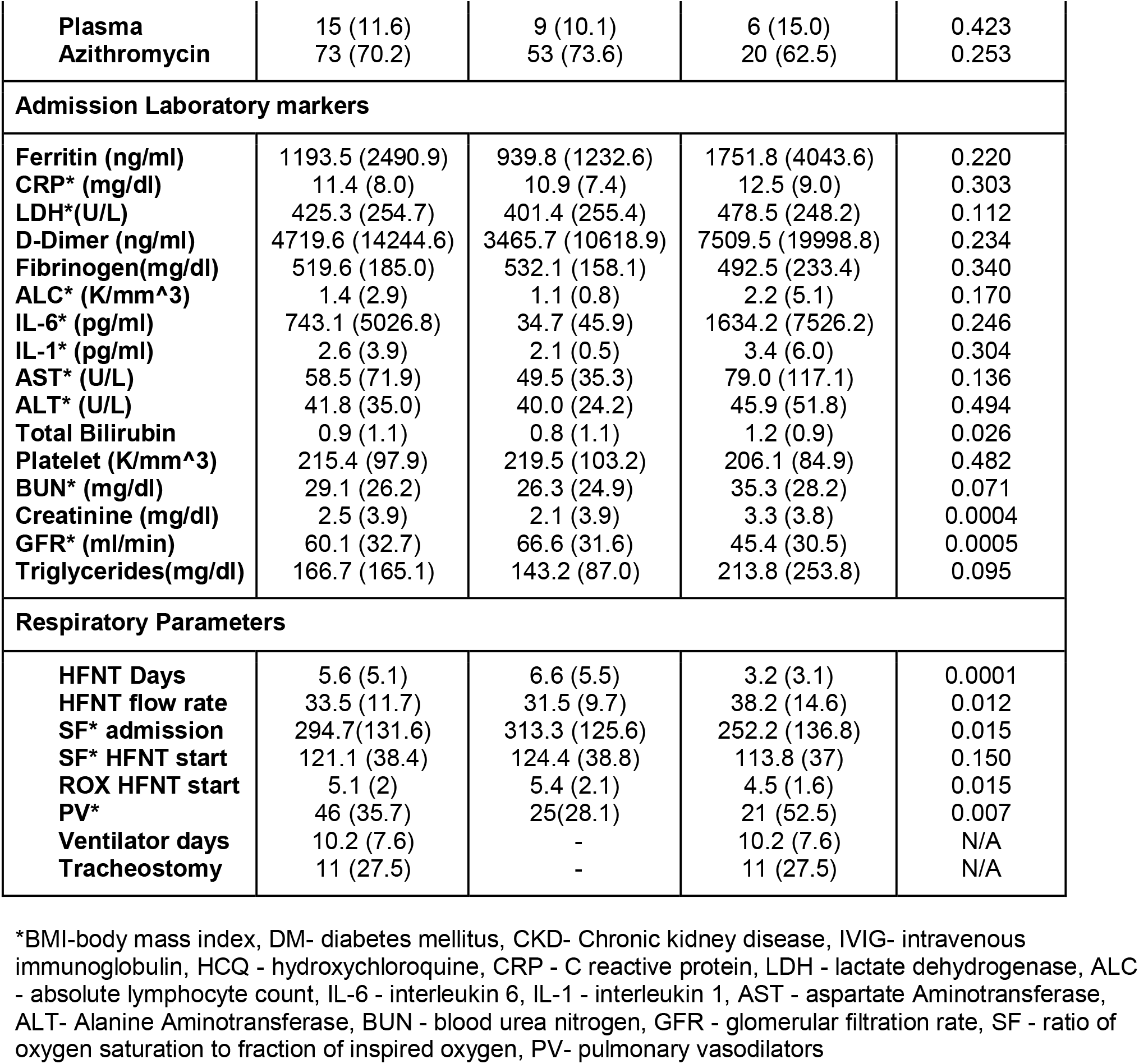
Baseline Demographics comparing HFNT group with HFNT progressed to IMV group

### Treatments

Azithromycin (70.2%) and steroids (86%%) were the most frequently utilized therapies. Immunomodulator therapy including sarilumab, anakinra, IVIG and tocilizumab were the next most commonly used therapies. There was a higher usage of gimsilumab, hydroxychloroquine, IVIG, tocilizumab and etoposide in the intubation group, while azithromycin was higher in the HFNT only group. Steroid usage and other immunomodulators were similar across the groups.

### Laboratory markers

Elevated inflammatory markers (i.e., ferritin, CRP, D-dimer, fibrinogen, LDH, IL-6), transaminitis and lymphopenia were observed in all patients. There was a trend towards higher inflammatory markers (i.e., ferritin, CRP, LDH, D-dimer, IL-6, IL-1), triglycerides and transaminases in the intubation group. Statistically significant higher creatinine and lower GFR were seen in the intubation group.

### Respiratory parameters

Mean admission S-F ratio was 294.7± 131.6 and was statistically different between the groups (313.3 ± 125.6 vs. 252.2 ±136.8). S-F ratio at high flow initiation was 121.1±38.4 overall, with no statistically significant differences in the groups (HFNT group 124.4± 38.8) vs intubation group (113.8±37). The mean corresponding P-F ratio at start of HFNT was ∼100.

Initial HFNT settings were 33.5±11.7 L/min of flow, while F_I_O_2_ was 84.1 % ±20.3. The intubation group had a statistically higher flow rate than the HFNT group. The average use of HFNT for our population was 5.6 days ±5.1. The minimum settings on HFNT were 10 L flow and F_I_O_2_ of 30%, while the maximum settings were 60 L and F_I_O_2_ of 100%. The major complication with use of HFNT was progression to IMV or NIV which was seen in 40 (31.0%) patients. Average ventilator days were 10.2±7.6 days. 10 (27.5%) patients received a tracheostomy. Overall, 46 patients required pulmonary vasodilators, with statistically higher usage in the intubation group.

### Outcomes

#### ROX index trends

Mean ROX index for the total cohort was 5.1 ±2.0 at HFNT initiation, 5.9±2.5, 6.9 ±3.9, 8.1±4.1 and 10.3±5.9 on day 1, 2, 3 and 5 respectively. The mean ROX index consistently improved from initiation to day 5 in the HFNT group, while staying constant in the intubation group (Figure 2). At each time interval, the ROX index was significantly higher in the HFNT group compared to the intubation group. The ROX change per day was also statistically different between the groups: (1.2±1.3) in the HFNT group vs. (−0.3

**Figure 2:**
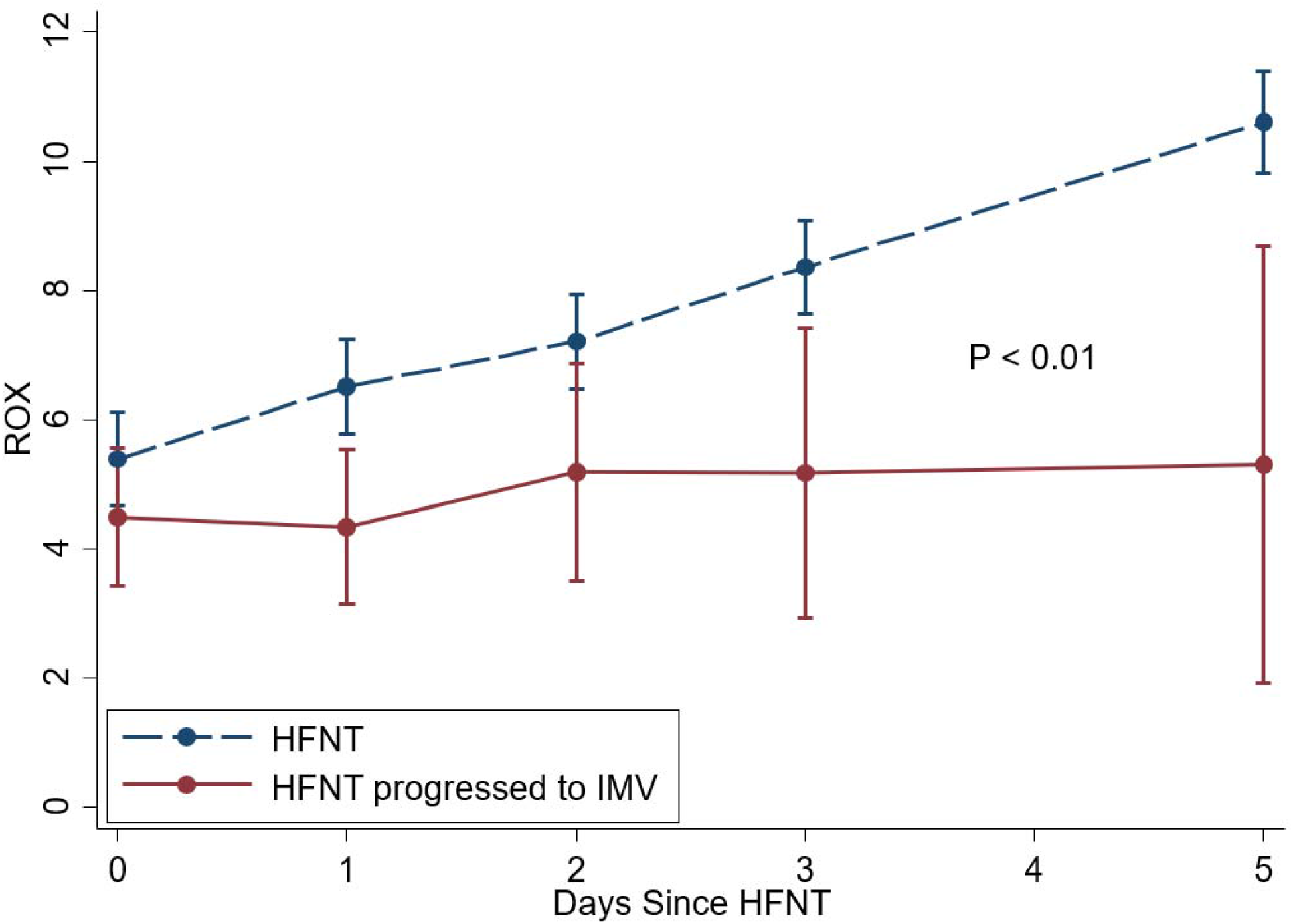
Average ROX index progression of HFNT group compared to HFNT progressed to IMV group.

±1.2) in the intubation group). ROX before intubation was the lowest at 3.4±1.0 (Table 2).

**Table 2:** ROX trends comparing HFNT group with HFNT progressed to IMV group

|  | N | Total | HFNT | HFNT progressed to IMV | p-value |
| --- | --- | --- | --- | --- | --- |
| ROX at HFNT initiation | 129 | 5.1 (2.0) | 5.4 (2.1) | 4.5 (1.6) | 0.015 |
| ROX at day 1 | 119 | 5.9 (2.5) | 6.5 (2.4) | 4.3 (1.8) | 0.0001 |
| ROX at day 2 | 101 | 6.9 (3.1) | 7.2 (3.2) | 5.2 (2.1) | 0.017 |
| ROX at day 3 | 98 | 8.1 (4.1) | 8.4 (4.2) | 5.2 (1.9) | 0.0006 |
| ROX at day 5 | 78 | 10.3 (5.9) | 10.6 (5.9) | 5.3 (2.0) | 0.078 |
| ROX at IMV | 40 | 3.4 (1.0) |  | 3.4 (1.0) | N/A |
| Mean ROX change per 24 hours | 129 | 0.7 (1.5) | 1.2 (1.3) | -0.3 (1.2) | 0.0001 |
| Median ROX change per 24 hours (IQR) | 129 | 0.5 (0-1.5) | 1.2 (0.3 -1.7) | 0 (-0.5 - 0.1) | 0.0001 |

#### Secondary Outcomes

Overall, mortality at our institution was 6.06 % for patients positive for COVID-19 infection. However, in this cohort of severe hypoxemic respiratory failure, our mortality was 22.5%, with 11.2% in the HFNT group and 47.5% in the intubation group. Figure 3 shows the Kaplan Meir curve between two groups for survival. Of the 10 deaths in the HFNT group, 6 patients were in hospice care while the remaining were Do not resuscitate/intubate (DNR/DNI). Average LOS was statistically higher in the intubation group (11.1 days in the HFNT group vs. 19.5 days in the intubation group) (Table 3).

**Table 3:** Other Outcomes comparing HFNT group with HFNT progressed to IMV group

|  |  | Total (N=129) | HFNT only (N=89) | HFNT progressed to IMV (N=40) | P-value |
| --- | --- | --- | --- | --- | --- |
| **Days to IMV** | **Mean**<br>**Median** | 2.5 (3.3)<br>1 (1.0 -3.0) | - | 2.5 (3.3)<br>1 (1.0 - 3.0) | N/A<br>N/A |
| **Mortality** |  | 29 (22.5) | 10 (11.2) | 19 (47.5) | 0.0001 |
| **LOS** |  | 14.0 (8.0) | 11.1 (4.7) | 19.5 (9.9) | <0.0001 |
| **HAP/VAP** |  | 11 (8.6) | 1 (1.1) | 10 (25.0) | <0.0001 |

\*LOS- length of stay, HAP/VAP - hospital acquired pneumonia/ventilator acquired pneumonia

**Figure 3:**
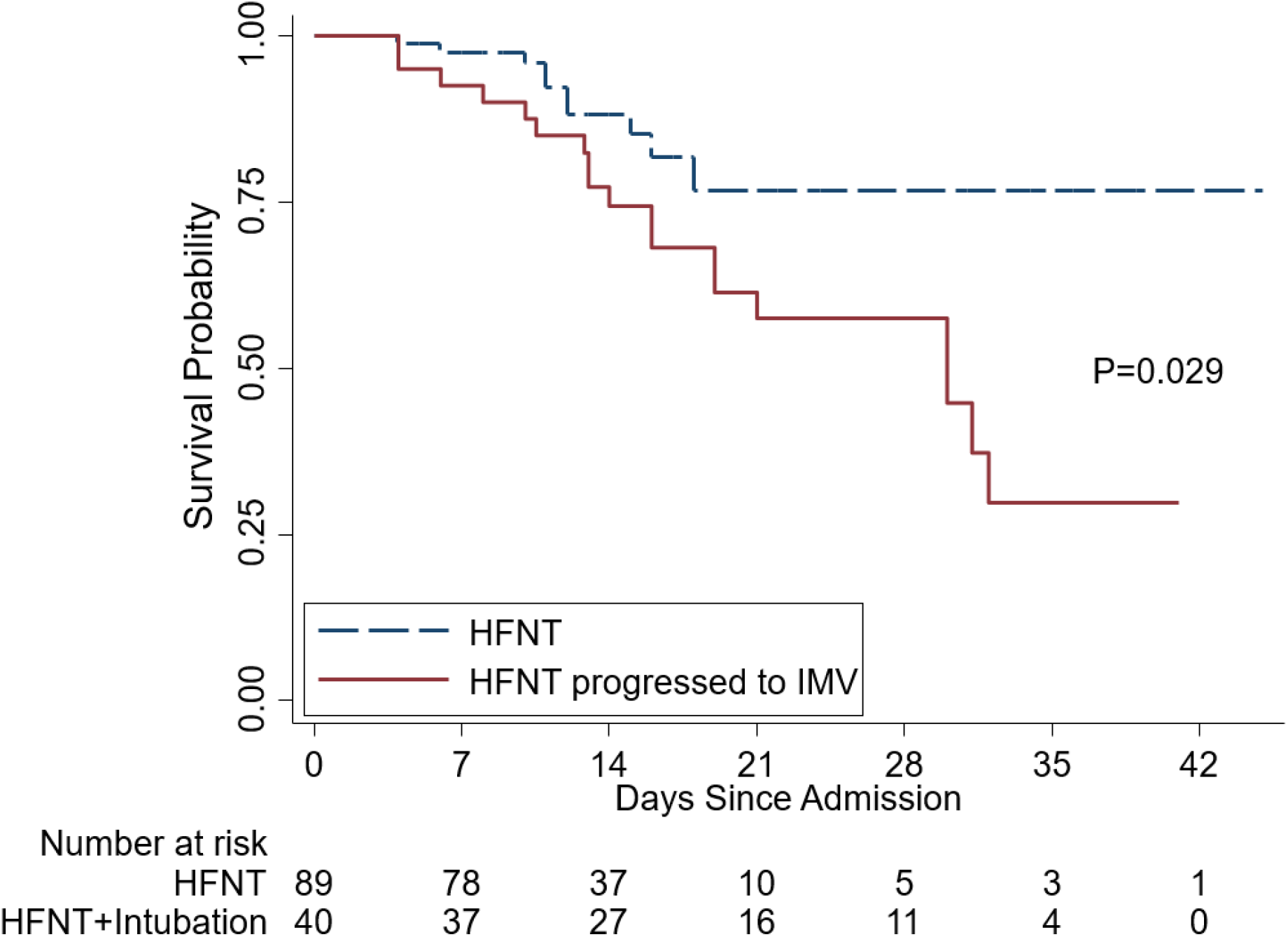
Kaplan-Meier Comparing survival in HFNT group and HFNT progressed to IMV group.

The overall incidence of hospital acquired pneumonia was significantly higher in the intubation group (25% vs 1.1%, p= 0.0001)

### Prediction Model

At initiation of HFNC, a ROX of < 5 was predictive of intubation (OR = 2.137, p=0.051). Any change in ROX of less than or equal to zero after HFNT initiation over 24 hours was also predictive of intubation (OR = 14.67, p <0.0001). A decrease in ROX by 1 over 24 hours regardless of ROX index value was strongly predictive of intubation (OR = 5, p <0.0001) (Table 4). Figure 4 shows intubation free survival based on ROX change (<=0 versus >0) per 24 hours. In the univariate analysis, smoking, history of malignancy, admission LDH > 500, peak D-dimer greater than 4000, peak Ferritin > 1000, Peak CRP >= 10, peak LDH > 500, ROX decrease as described above, admission triglycerides > 200, and a glomerular filtration rate < 60 were all predictive of intubation (See supplementary Table 1). In a multivariate model, unchanged and/or decreased ROX over 24 hours, peak D-dimer greater than 4000 and GFR less than 60 ml/min were predictive of intubation (Table 4). Figure 5 and 6 show the receiver operator curve for ROX change over 24 hours (ROC = 0.77) and the multivariate model respectively (ROC = 0.86).

**Table 4:** Logistic Regression Model Predicting Need for IMV

| Variable | Odds Ratio | P-value |
| --- | --- | --- |
| ROX at HFNT initiation<br>=<5<br>>5 | 2.137<br>1 | 0.0517 |
| ΔROX from baseline (any 24-hr period)<br>Decreased by 1<br>Increased by 1 | 5<br>1 | 0.0001 |
| ΔROX change per day<br>=< 0<br>> 0 | 14.671<br>1 | 0.0001 |
| Pulmonary Vasodilators<br>Yes<br>No | 2.83<br>1 | 0.0084 |
| **Final Multivariate Model** |  |  |
| ΔROX change per day (=< 0 vs > 0) | 13.17 | 0.001 |
| Peak D-dimer (>=4000 vs <4000) | 4.47 | 0.0026 |
| GFR (<=60 vs >60) | 3.29 | 0.0163 |

\*Univariate model in supplementary

**Figure 4:**
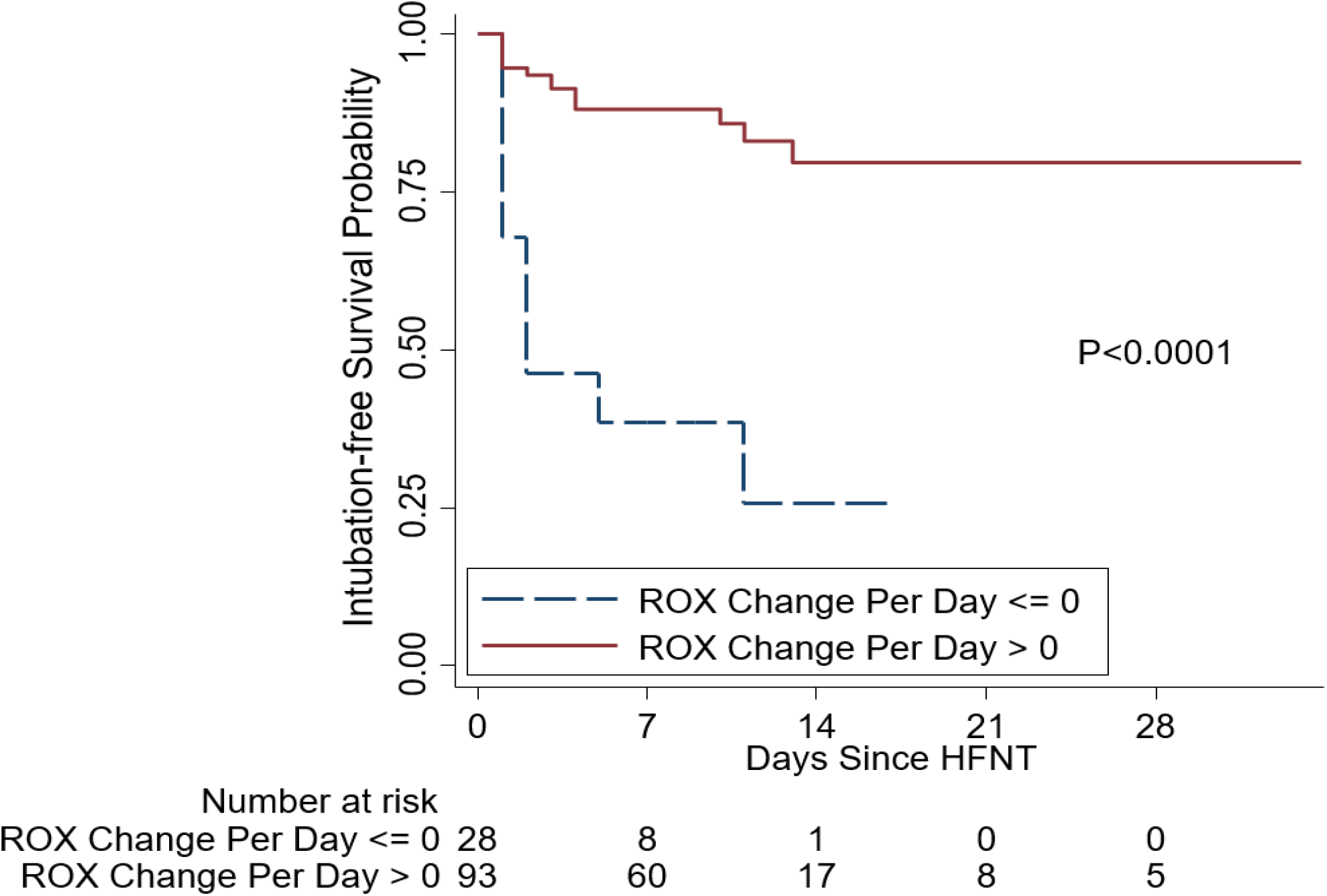
Kaplan-Meir showing intubation Free Survival probability by ROX change per 24 hours.

**Figure 5:**
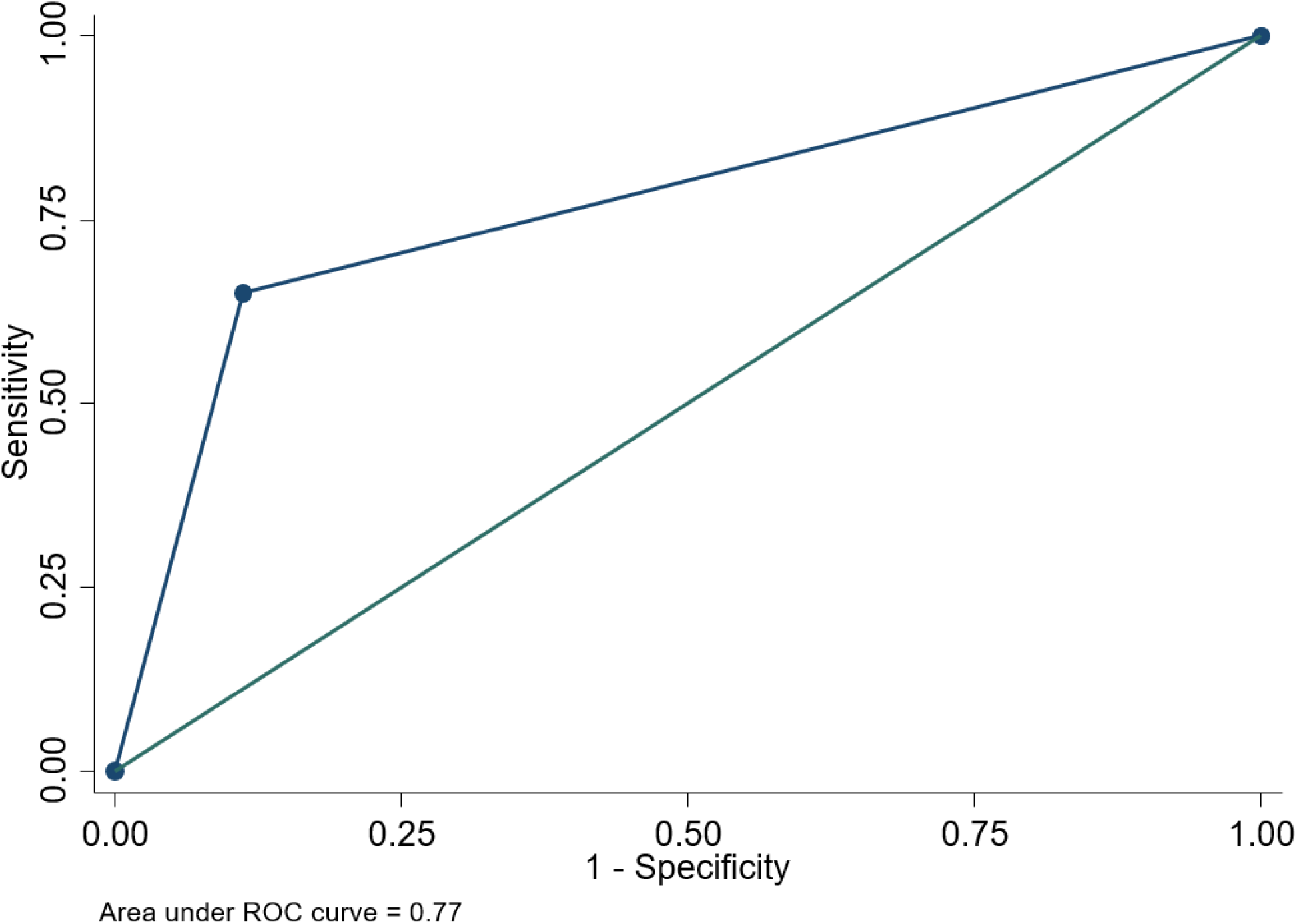
ROC predicting need for IMV using ΔROX per 24 hours.

**Figure 6:**
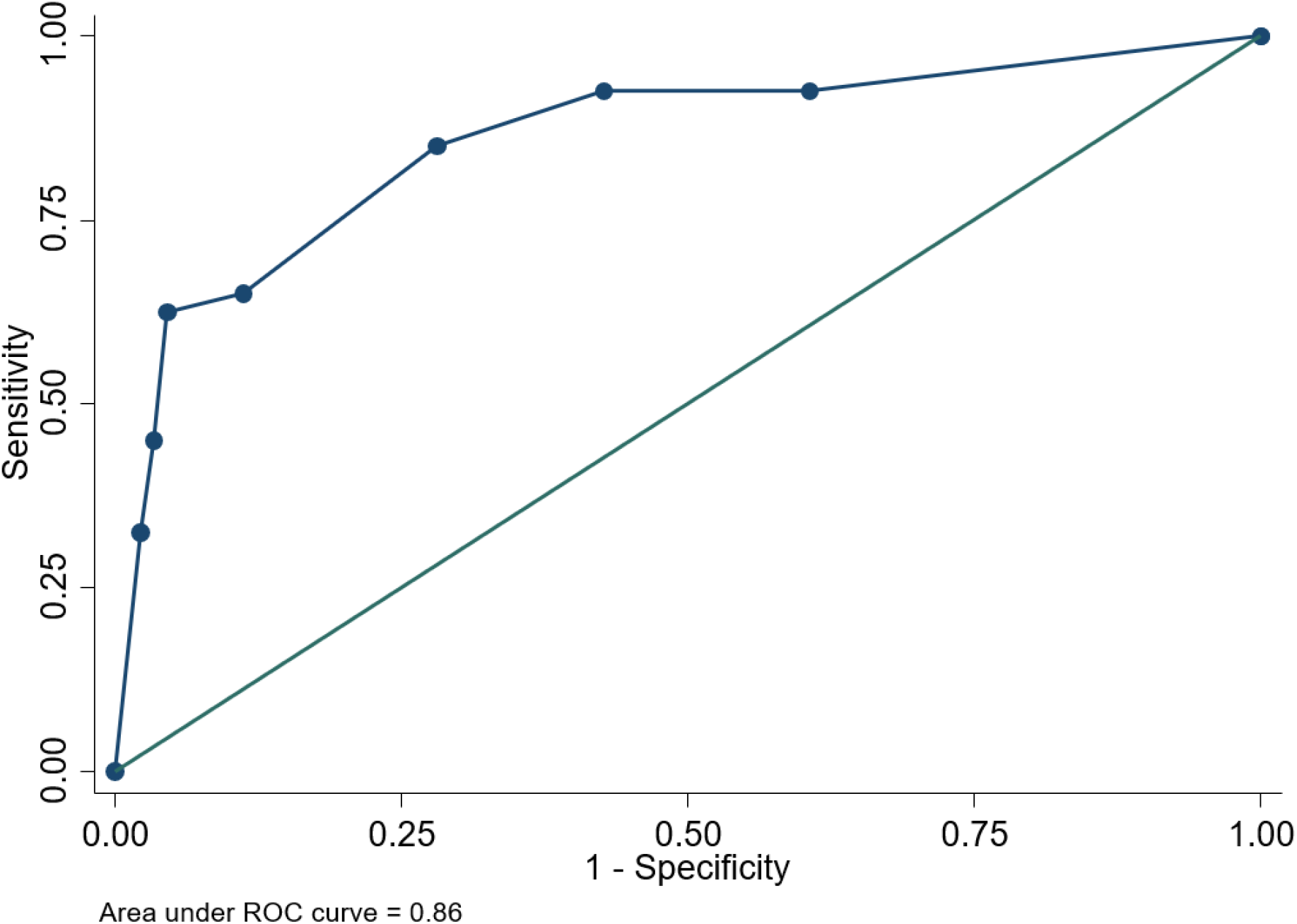
ROC of multivariate model of ΔROX, D-Dimer and GFR to predict need for IMV.

## Discussion

In this retrospective review of patients with acute hypoxemic respiratory failure secondary to COVID-19 pneumonia, 129 patients were initially treated with HFNT. Out of this cohort, 89 patients remained on HFNT while 40 patients eventually required IMV.

The 89 patients successfully treated with HFNT as a bridge to recovery had a significant improvement in ROX from initiation of HFNT at all recorded time points. In contrast, the ROX score for patients that ultimately required intubation remained steady or decreased over time.

A decrease in ROX score of 1 over any 24-hour period increases the likelihood of intubation 5-fold (OR = 5, p < 0.0001), regardless of ROX value at HFNT initiation. When compared to the baseline ROX score at HFNT initiation, failure of the ROX score to improve (ΔROX ≤ 0) increases the likelihood of intubation by a factor of 15 (OR = 14.67, p<0.0001). Lastly, a combination of change in ROX index, GFR<60 and peak D-dimer >4000 were even more predictive of intubation. Mortality and LOS in the HFNT group was significantly lower than the intubation group and the incidence of hospital-acquired pneumonia was higher in the intubation group. There were no associated deaths peri-intubation despite the presence of significant hypoxemia. There were no reported cases of failure to intubate resulting in adverse outcome. Overall, the intubation group had a higher incidence of lung disease, chronic kidney disease, smoking, and malignancy.

HFNT is an important oxygen delivery modality that can help reduce intubation as seen by our overall institution intubation rate of 10%, significantly lower than the reported literature.^4,6,7^ Moreover, there may be a survival benefit with HFNT therapy in COVID-19 as seen in prior acute hypoxemic respiratory failure studies.^13, 25^ Despite our patient population having a higher incidence of lung disease and nicotine exposure than that reported in previous studies, the mortality rate was 22%, which is lower than prior reports.^4,6,11^.

Gattinoni and colleagues proposed that COVID-19 patients fall into two distinct groups or phenotypes. The “Type L” or “non-ARDS Type 1” phenotype has low elastance and high compliance. These patients often present with profound hypoxemia and low lung recruitability. As opposed to this the “Type H” or “ARDS Type 2” phenotype have high elastance and low compliance, requiring traditional management strategies of higher PEEP and lower tidal volumes.^26,27^ A significant number of COVID-19 patients present with silent hypoxemia. As HFNT provides a modest PEEP effect (i.e. 3-5 cmH_2_O at flow rates of 30-50 LPM with mouth closed)^28^ patients with predominant Type L physiology may benefit from the oxygenation support that HFNC can provide noninvasively. HFNT also leads to a high oxygen reservoir by reducing anatomical dead space in the nasopharynx.^29^ Often, higher tidal volumes are employed in type-L phenotype which can lead to ventilator associated lung injury (VILI). VILI can cause inflammatory cytokine release in ARDS patients, including IL-6, both in critically ill humans.^30, 31^ IL-6 in particular is one of the pathologic mechanisms for lung injury in COVID-19.^32,33^ Thus, use of HFNT should not be overlooked in patients with severe COVID-19 respiratory failure.

Patient self-induced Lung injury (P-SILI) has been cited as a theoretical contraindication to noninvasive methods of oxygenation. To date however, P-SILI remains a conceptual model concept compared to VILI.^34,35^

Optimal timing of IMV remains a point of debate, especially in patients previously supported with noninvasive forms of oxygen support, especially with regards to COVID-19. Based on our results, any decrease in ROX index over a 24-hour period from baseline ROX at HFNT initiation is a strong predictor of intubation, irrespective of total number of HFNT days. We choose to design ROX change as ≤ 0 vs. >0 for ease of use in the acute care setting.

Roca et al previously used a ROX index of < 4.8 at 12 hours to successfully identify patients with high risk for intubation amongst a cohort of 191 patients treated with HFNC for acute hypoxemic respiratory failure secondary to pneumonia.^23,36^ Our analysis further validates their findings in the setting of viral pneumonia as opposed to predominantly bacterial pneumonia as was reported in their study. Our receiver operator analysis yielded similar results to initial studies. Thus, using serial measurements, we can easily identify patients on HFNT therapy in whom IMV should be considered based on changes in ROX.^37^

Theoretically, the ROX can easily identify patients shifting from L-phenotype to H-phenotype (lower SF ratios and higher respiratory drive), thus minimizing subsequent risks of P-SILI. Another advantage of using the ROX index is its noninvasive nature based on readily available clinical parameters. The ROX index can be calculated remotely, thus preserving personal protective equipment and limiting healthcare exposure. When combined with a decreasing ROX index, a GFR <60 and D-dimer >4000 stratifies high risk patients with increased accuracy. Kidney dysfunction makes patients susceptible to even small fluid shifts, thus worsening hypoxemia. D-dimer > 4000 might possibly be a sign of micro thrombi in pulmonary circulation described in COVID-19.^38^

Viral transmission through aerosolization by non-invasive forms of oxygenation such as HFNT remains controversial and is much debated. During the SARS outbreak in 2003, transmission to healthcare workers was reported from only 8% of HFNT patients.^39^ This was demonstrated in further studies that proved that bacterial environmental contamination was not increased in the setting of HFNT use.^40^ An in-vitro study mimicking clinical scenarios including HFNT with mannequins only revealed proximal dispersion of secretions to the face and nasal cannula itself.^41,42^ A recent study with healthy volunteers wearing high-flow nasal cannulas at both 30 L/min and 60 L/min of gas flow did not report variable aerosolization of particles between 10-10,000 nm, regardless of coughing, when compared with patients on room air or oxygen via regular nasal cannula.^43^ At an institution with dedicated COVID-19 wards, only 1 of 80 staff members in our department had suspicion of health care transmission while directly caring for COVID-19 patients, thus reemphasizing that HFNT did not present an increased risk of healthcare transmission.

Our study has several strengths. It is the largest reported cohort utilizing HFNT in COVID-19 thus far. The ROX index was able to successfully predict bridge to recovery or progression to IMV without demonstrable adverse effect from delaying implementation of mechanical ventilation. In a high-risk, urban population with multiple comorbidities, use of HFNT resulted in a lower rate of intubation, and suggests a possible mortality benefit while maintaining a low risk of healthcare transmission.

Our study has several limitations. First, it is a retrospective review, thus making it susceptible to unintended biases. Developing a prospective study during a pandemic situation was impractical. Secondly, although this is the largest HFNT study, the total N is limited and representative of a singles center’s experience. Lastly, we were unable to provide consistent details on the presence and degree of hypercapnia for our cohort due to our institutional policy to minimize staff exposure to COVID-19 infection.

In conclusion, the ROX index provides an accurate risk stratification tool in patients with moderate to severe hypoxemic respiratory failure secondary to COVID-19 pneumonia. HFNT can be safely and successfully implemented while utilizing the ROX index to predict the need for IMV. Monitoring ROX trends may allow clinicians to avoid any significant delays in escalating the level of care or implementing IMV. Use of HFNT not only reduces intubation rates, but also has the potential to reduce mortality and morbidity associated with IMV.

## Data Availability

Data available upon request

## Acknowledgements

Maulin Patel will be the corresponding author and guarantor for the manuscript. Maulin Patel, Matthew Gordon, Junad Chowdhury and Gerard J Criner formulated the overall study design. Huaqing Zhao, Nicole Patlakh, Maulin Patel, Andrew Gangemi, Robert Marron, Junad Chowdhury, Nicole Mills, Zachariah Dorey-Stein, Ibraheem Yousef, Lauren Tragesser, Julie Giurintano assisted in data collection, consolidation and analysis. Maulin Patel, Junad Chowdhury, Parth Rali, Rohit Gupta, Gilbert D’Alonzo and Matthew Gordon drafted the manuscript. Gerard J Criner and Matthew Gordon revised and reviewed the Manuscript

